# Effectiveness of Influenza-Prevention Interventions among Healthcare Workers: A Systematic Review and Meta-Analysis of Health Outcomes

**DOI:** 10.1101/2025.06.23.25330051

**Authors:** Ahmed Al Tamimi, Saoud Al Tamimi, Khalifa Al Seiari, Rami H AlRifai, Susannah Fleming

## Abstract

**Background:** Although studies do not provide conclusive evidence of their benefits, influenza vaccinations and face masks are recommended and even mandated to prevent influenza infections in healthcare workers (HCWs).

**Objectives:** To summarise the latest evidence on the effectiveness of influenza prevention interventions in HCWs.

**Methods:** We systematically searched PubMed, Scopus and Google Scholar for RCTs, cohorts and cross-sectional studies published in English up to 30 September 2024. All studies comparing groups of HCWs with and without intervention/exposure were included. Three reviewers independently selected articles and extracted data. Estimates were pooled using random-effects meta-analyses. Pooled analyses were conducted on outcomes including laboratory-confirmed influenza (LCI), influenza-like illness (ILI), and work absenteeism.

**Main results:** Twenty-one articles met eligibility criteria. For influenza vaccine, three articles were RCTs, twelve were cohort and three were cross-sectional studies; and for face masks, there was one RCT, one cohort and one cross-sectional study. The pooled results showed an insignificant effect of influenza vaccine and face masks on the incidence of laboratory-confirmed influenza (RR= 0.68, 95% CI: 0.36–1.27) and ILI (RR= 0.23, 95% CI: 0.03–1.68). A subgroup analysis showed that vaccination significantly reduced the incidence of LCI in small samples (<30 participants), but not in large samples. In addition, influenza vaccination was not associated with reducing the incidence of ILIs (RR= 1.04, 95% CI: 0.84–1.29). However, it significantly reduced work absenteeism (SMD= 0.87, 95% CI: 0.81–0.94). There is insufficient data to assess the effects of wearing a face mask on ILI or workplace absenteeism.

**Conclusion:** Our findings did not provide conclusive evidence for the effectiveness of influenza vaccination or face masks in reducing influenza infections. Influenza vaccination had a significant benefit in reducing absenteeism in HCWs by 17%. As HCWs play a central role in patient care, it is crucial to ensure the safety and protection of patients. Therefore, understanding the clinical need for influenza protection while applying other practical measures such as hand hygiene and other personal protective equipment is essential. High-quality RCTs are needed to evaluate the final impact of these protective measures in different clinical settings and parts of the world.

## Background

Seasonal flu (influenza) is caused by viruses that spread mainly through droplets and contact with infected patients (1) Influenza causes severe illness in 3 to 5 million people and kills 290,000 to 600,000 people worldwide every year (2). It is often accompanied by fever, runny nose, sore throat, muscle aches, headache, cough and fatigue. These symptoms can be caused primarily by influenza A and B viruses, the most common influenza-like illness (ILI) triggers. However, other viruses can also cause ILI, including respiratory syncytial viruses, rhinoviruses, adenoviruses, parainfluenza viruses and human coronaviruses (3). The course of influenza is usually mild and self-limiting and is rarely fatal (4) but can be severe depending on various factors and conditions (e.g. age, immune status and comorbidity) (2). In addition, influenza can pose a significant health risk to vulnerable groups as it can develop into viral pneumonia or subsequent bacterial infections (5). Other complications of infection include acute respiratory distress syndrome, meningitis, encephalitis and exacerbation of pre-existing health problems such as asthma and cardiovascular disease (6).

Although influenza affects 10-20% of the population worldwide each year (7), hospitals and other healthcare facilities can be overwhelmed with sick patients during an outbreak, putting healthcare workers at increased risk of infection and transmission (8). As droplets mainly transmit the influenza virus, frequent hand washing and covering the mouth and nose when coughing and sneezing should at least theoretically protect against infection. This may explain why the European Centre for Disease Prevention and Control considers these measures to be adequate personal protection when caring for a patient with influenza infection (5). However, the effectiveness of these protective measures remains controversial (9). Therefore, annual influenza vaccination is the most effective means of preventing influenza and influenza-related complications, especially for high-risk groups (5). There are two types of influenza vaccines: the inactivated influenza vaccine (IIV) and the live attenuated influenza vaccine (LAIV), with no influenza vaccine being preferred over the other (5). As the influenza virus is susceptible to antigenic changes, influenza vaccines are only effective if there is an antigenic match between the vaccine and the circulating virus strains (2). The WHO’s Global Influenza Surveillance and Response System (GISRS) works to reformulate the vaccine each flu season to match the circulating virus strains (2). Despite these efforts, the effectiveness of the vaccine has been in the same range in previous seasons, namely between 40% and 60% (10). In the 2022-2023 season, efficacy was 51% (95% CI: 33%-64%) in the 18-64 age group.

The World Health Organisation (WHO) considers influenza vaccination a priority for vulnerable people and healthcare workers (2). The CDC has also recommended seasonal vaccination for all healthcare workers who are at higher risk of contracting influenza and transmitting it to their patients (11). Several systematic reviews have already evaluated the effectiveness of influenza vaccines in different groups of high-risk individuals. For example, a recent systematic review found that vaccination of older adults living in care facilities plays a protective role (12). On the other hand, few reviews have examined the impact of influenza vaccines on outcomes in healthcare workers. Ng et al. 2011 concluded in very limited studies (three RCTs) that there was insufficient evidence of the effectiveness of influenza vaccines (13). A Cochrane review has concluded that offering influenza vaccination to healthcare workers caring for people aged 60 years or older has little or no effect on laboratory-detected influenza (14). More recently, Li et al. 2021 conducted a systematic review and meta-analysis to assess the impact of influenza vaccination on outcomes in healthcare workers. Their analysis showed that influenza vaccination helped to reduce the incidence of laboratory-confirmed influenza (LCI) in vaccinated healthcare workers, absenteeism rates, and workdays lost (15).

Our review aims to understand the effect of influenza vaccination and face masks on reducing influenza incidence in healthcare workers so that appropriate infection control measures can be taken to reduce influenza transmission in hospitals and improve staff productivity. In this review, we investigate whether vaccination or the use of face masks for healthcare workers affects the incidence of (i) laboratory-confirmed influenza, (ii) influenza-like illness, and (iii) days of absenteeism among healthcare workers.

## Methods

This systematic review and meta-analysis followed the Cochrane guidelines (16) and the Preferred Reporting Items for Systematic Reviews and Meta-Analysis (PRISMA) guidelines (17).

### Study Design

Systematically applying the PI/ECO (population, intervention/exposure, comparison, and outcome) approach, this paper combines interventional and observational studies of influenza prevention measures to clarify controversies about the conflicting claims of their effectiveness. Studies were included if they were RCTs, cohort, or cross-sectional studies. For more information about PI/ECO, refer to the supplementary appendix A1.

### Search Strategy

We searched three databases, PubMed, Scopus, and Google Scholar, and used the following combinations or search terms to search the databases: “influenza vaccine”,” “face mask”,” “healthcare workers” and “effectiveness” in the title, abstract or keyword field. The full list of search terms and the search strategy can be found in supplementary appendix A2. The reference lists of relevant articles were also hand-searched for eligible studies. The database search was conducted on 30 September 2024 with no time restriction on the publication date.

Studies that provided results for multiple years/seasons were included as separate entries based on the respective year/season. In addition, only studies that compared intervention arms that met our inclusion criteria were included. No studies were excluded due to the high risk of bias. We excluded case studies or studies that were not conducted in hospitals or medical clinics, as well as studies published in languages other than English.

### Citations screening and identifying eligible studies

Three reviewers (ST, KS, and AT) independently reviewed the titles and abstracts to assess the literature’s eligibility. The full texts of all studies that were or could be considered for the study and those included in a previously published systematic review were thoroughly screened and assessed for eligibility. All discrepancies were resolved through discussion and consensus.

### Data extraction

The data of the eligible studies were extracted independently by three reviewers (ST, KS and AT). The reviewers evaluated all full-text articles and assessed the completeness of the data and the risk of bias. A structured data extraction form was created using Excel (Microsoft® Excel® for Microsoft 365) to ensure consistency of the data extraction process. Any discrepancies were identified and clarified in follow-up meetings to reach a consensus. Data extracted included study characteristics (first author name, year and country of publication), study design (RCT, cohort and cross-sectional study), intervention and comparator information (type of intervention/comparator), number of HCWs with incidence of outcomes of interest and total number of participants in the intervention/exposure group and in the control/non-exposure group. Regarding absenteeism, the mean, standard deviation and number of participants for lost working days were extracted.

### Statistical analysis

The results were presented descriptively. The association between each intervention/exposure and the corresponding control arm was presented as the risk ratio (RR) for laboratory-confirmed influenza and the incidence of ILI. The mean difference (MD) was used for the number of days lost from work. To account for expected heterogeneity between studies due to differences in vaccine types, laboratory methods (e.g. serology, rapid influenza diagnostic test (RIDT)) and population characteristics, we considered that a fixed-effects model would not assume a common true effect for all studies. Therefore, we presented the pooled estimate using DerSimonian and Laird’s method (random effects) to optimally capture the broader distribution of effect sizes and reflect both within-study and between-study variability, consistent with the uncertainty inherent in vaccine effectiveness data. Similarly, we pooled the mask studies using a random effects model because of differences in intervention definition, methodology (i.e., survey, self-report, or clinic visit), and outcome assessment.

The significance level was set at 0.05. The meta-analysis was performed using Stata software (version IC 14.2; Stata Corp, University Station, TX).

### Quality and risk of bias assessment

We used the Risk of Bias Tool developed by the Cochrane Group **(18)** and the Newcastle-Ottawa Scale (NOS) **(19)** to assess the quality of RCTs and non-RCTs, respectively. We categorised the quality of evidence for cohort and cross-sectional studies as high, moderate and low based on the NOS scores for selection, comparability and outcomes. The quality assessment of the included articles was performed independently by three reviewers (ST, KS and AT). Disagreements were resolved by discussion and consensus between the reviewers; the average score for each item was considered in the absence of consensus.

### Heterogeneity

This review assessed potential statistical heterogeneity using the I^2^ test statistic. Heterogeneity was categorised as insignificant if the I^2^ value was below 30%, while I^2^ values of 30% to 50% indicated moderate heterogeneity, I^2^ values of 50% to 75% indicated substantial heterogeneity, and I^2^ values of 75% to 100% indicated considerable heterogeneity, in accordance with Cochrane methodology **(20)**.

### Sensitivity and subgroup analyses

We conducted sensitivity analyses by excluding old studies conducted before 2010 and before 2015 to assess their impact on our analyses. We conducted pre-specified subgroup analyses to account for heterogeneity by stratifying by study design (RCT, cohort studies or cross-sectional studies) and study size based on the total number of participants: small (less than 30 participants), medium (30 to 200 participants) and large (more than 200 participants).

Small but well-conducted studies can provide a reliable estimate of the outcome. However, the sample size is directly related to the statistical power of study, studies with fewer than 30 participants are often underpowered, which increases the risk of type II error. Setting <30 as “small” corresponds to the widely accepted thresholds for minimum power.

A higher sample size (200 versus 100) was chosen to have a more reliable and stable estimates with narrower confidence intervals, which is helpful for investigating how study size contributes to heterogeneity. Studies with 100 participants may still have underpowered estimates, especially when conducting subgroup analyses. By setting the threshold at 200, we ensure that the studies are more likely to provide reliable and stable estimates with narrower confidence intervals.

### Assessing the publication bias

The potential for publication bias was assessed using the Funnel plots and the Egger test.

## Results

### Database search

A total of 1,102 studies were identified using the predefined search strategy. After eliminating duplicates, 827 citations were selected for title and abstract screening. We excluded 787 studies because they did not fulfil the eligibility criteria and added two studies from the reference list. Eventually, twenty-one studies that were found eligible were subjected to data extraction (Supplementary appendix B1). The study selection process and the corresponding results are summarised in Figure 1.

**Figure 1.**
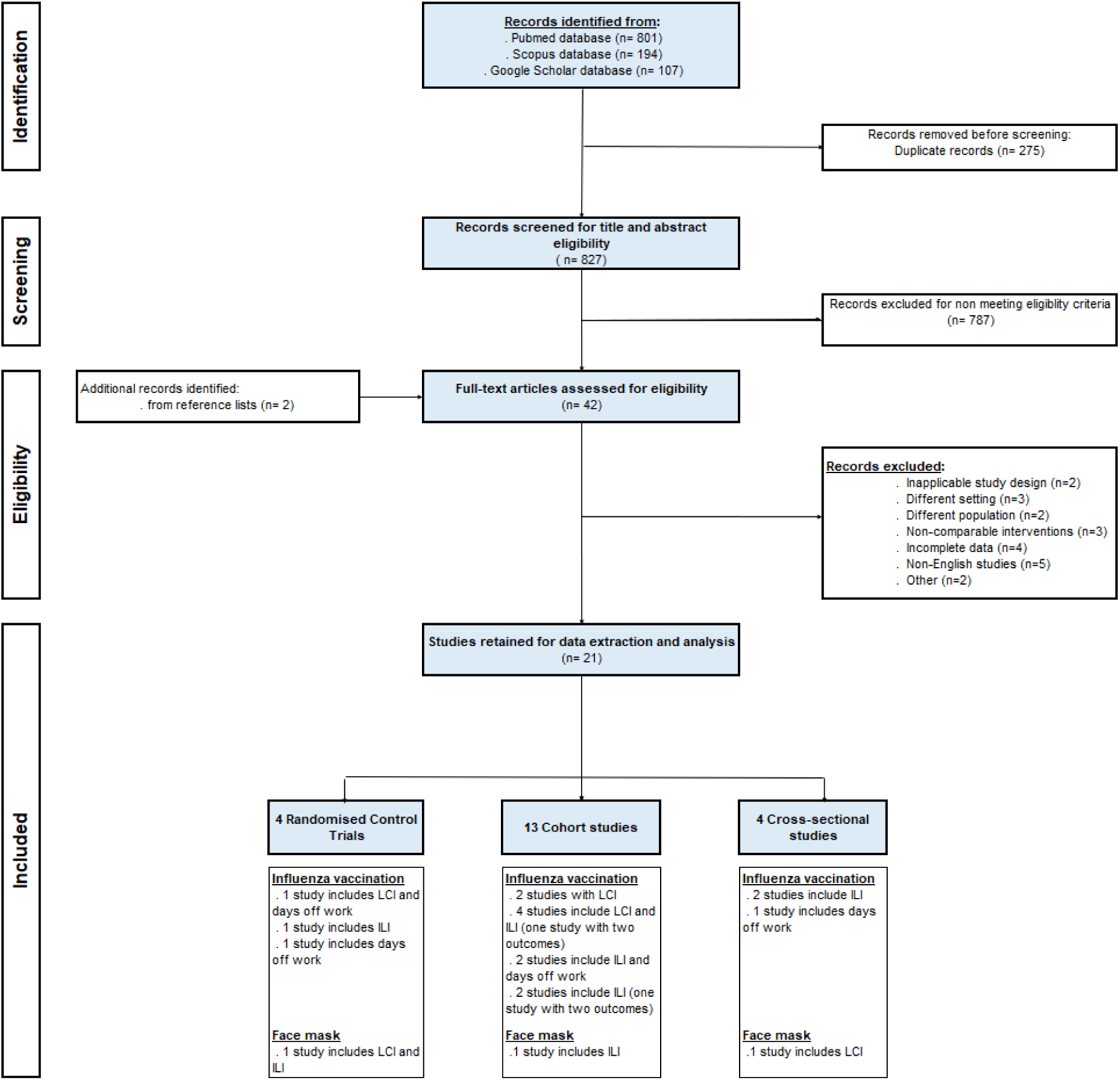
Flow-diagram of selecting eligible studies. LCI: Laboratory-confirmed influenza, ILI: Influenza-like illness.

### Scope of the review and characteristics of the included studies

Of the twenty-one included studies, four were RCTs, thirteen were cohort studies and four were cross-sectional studies. Table1 illustrates the distribution of these articles by type of outcome measured and intervention/exposure studied. For influenza vaccination, eighteen studies were conducted in twelve countries worldwide: four in Japan, three in Italy, two in the United States, and one each in Belgium, Canada, China, Finland, Hong Kong, Israel, Kenya, Singapore, and Taiwan. For the face mask intervention, three studies were conducted in Japan, the United States and Vietnam.

Three studies reported four outcomes for mask use (three reported LCI, of them, one also reported on ILI). These studies were conducted in Vietnam, the USA, and Japan. No study reported the number of days lost from work. Therefore, we only meta-analysed the studies that reported LCI.

For quality assessment results, two RCTs related to the vaccine intervention had a low risk of bias and one had a moderate rating, while the only RCT related to the mask intervention had a high risk of bias (Figures 2). The quality assessment of the cohort and cross-sectional studies is shown in Table 2.

**Figure 2.**
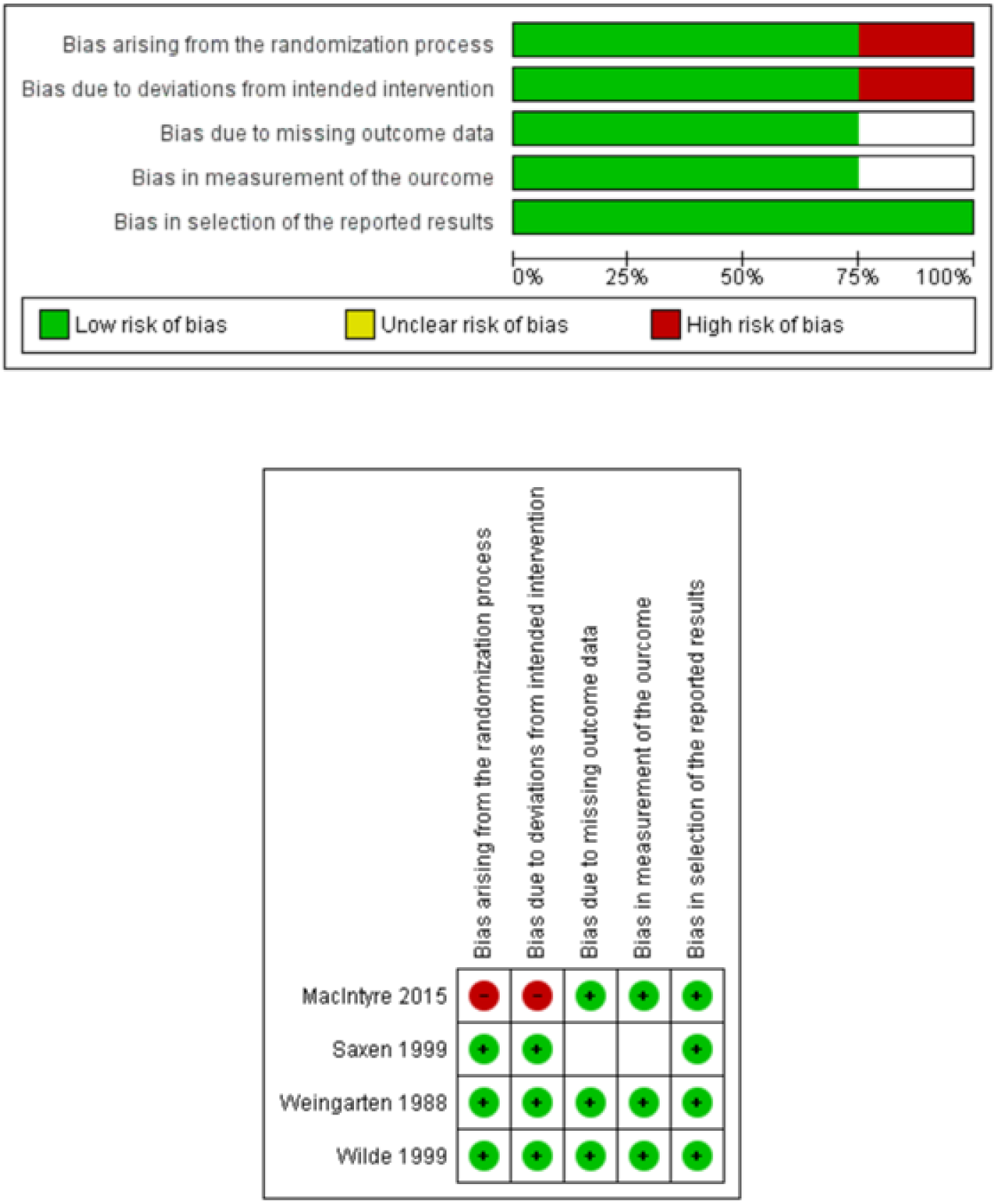
Quality assessment of the four randomised controlled trials, created using RevMan 5.4.1.

### Intervention/exposure – outcome pair

#### Influenza vaccine – Laboratory-Confirmed Influenza (LCI)

Seven studies investigated the incidence of influenza infections in vaccinated HCWs (Table 1); one RCT (Wilde JA, 1999) and six cohort studies (Atamna, 2016; Ishikane, 2016; Ito, 2006; Njuguna, 2013; Panatto, 2020; and Barbara, 2006), with 2-year data available in the latter study. The studies were conducted in North America, Europe, Africa, the Middle East and Far East Asia.

**Table 1.**
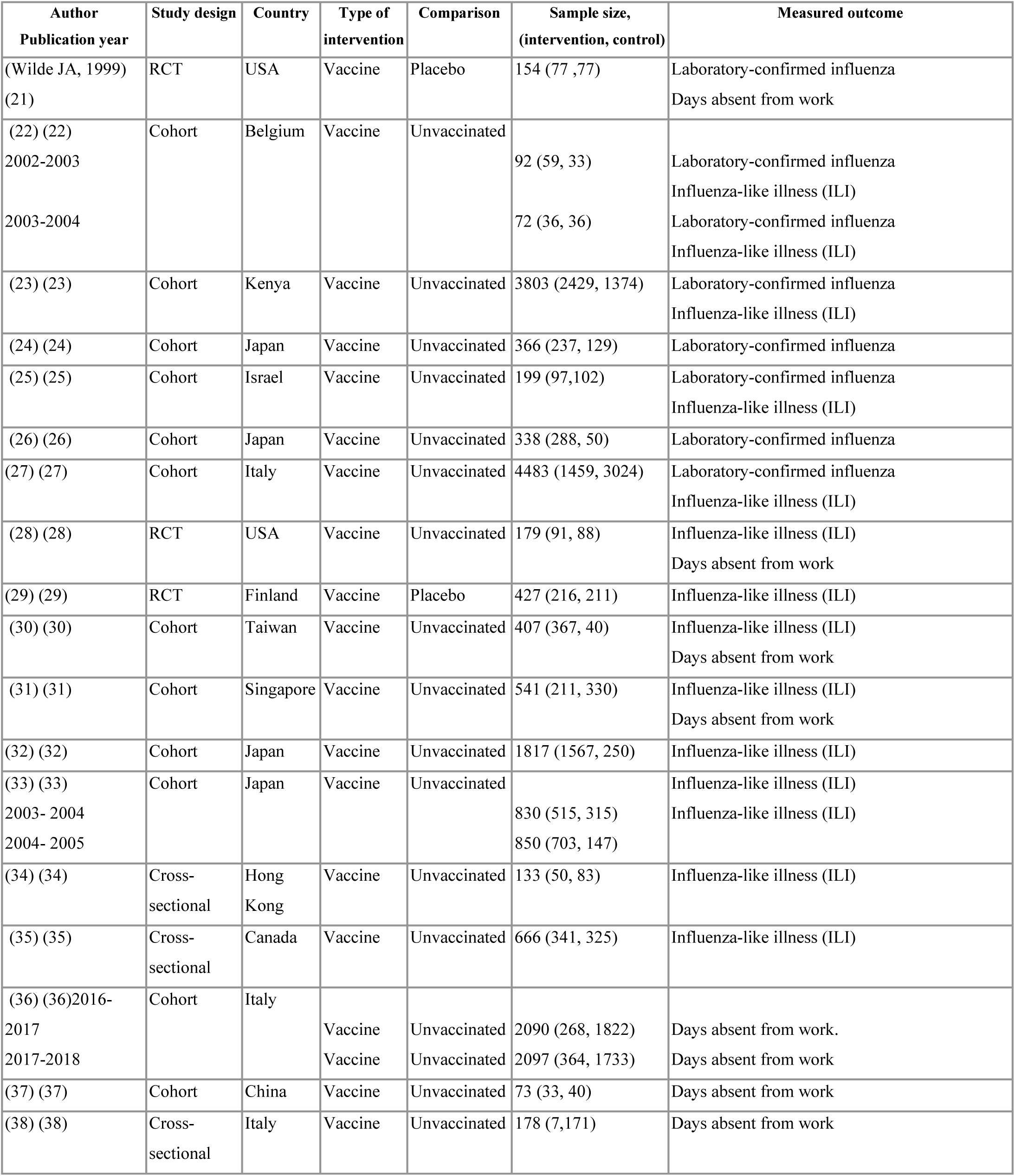
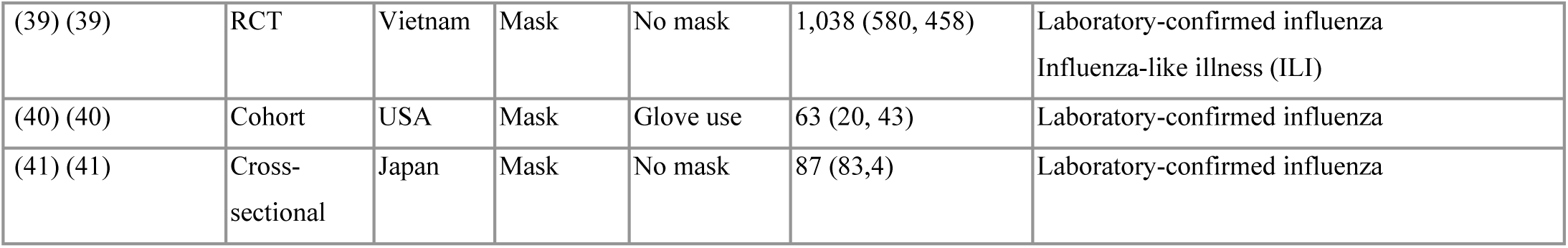
Baseline characteristics of the included studies.

**Table 2.**
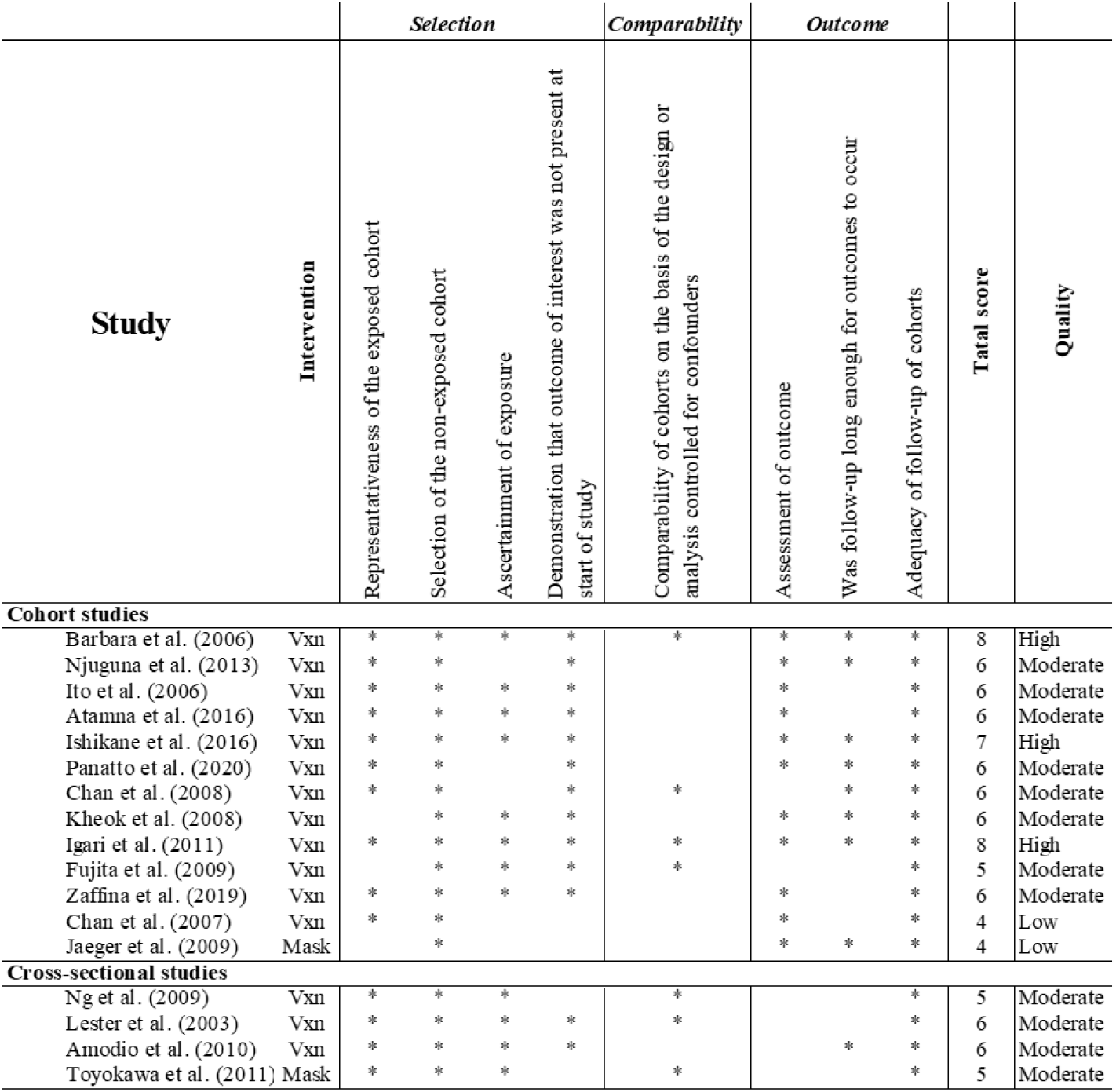
Risk of bias summary: review authors’ judgments for risk of bias items for each included cohort and cross-sectional study. *, the revised study met the criteria for this item, Vxn: vaccine.

The meta-analysis shows a statistically non-significant reduction in the incidence of laboratory-confirmed influenza among vaccinated compared to unvaccinated HCWs by 32% (pooled RR: 0.68, 95% CI: 0.36–1.27) (Figure 3). Cochran’s Q statistic showed substantial heterogeneity (I^2^: 72.7%, *p*-value = 0.001) in the included studies. A subgroup analysis by sample size showed a statistically significant reduction (RR: 0.41, 95% CI: 0.22 to 0.75, *p*-value = 0.004) in the risk of LCIs in medium sized studies compared to a non-significant effect size in the large studies (RR: 1.05, 95% CI: 0.50 to 2.18, *p*-value = 0.900 (Figure 4).

**Figure 3.**
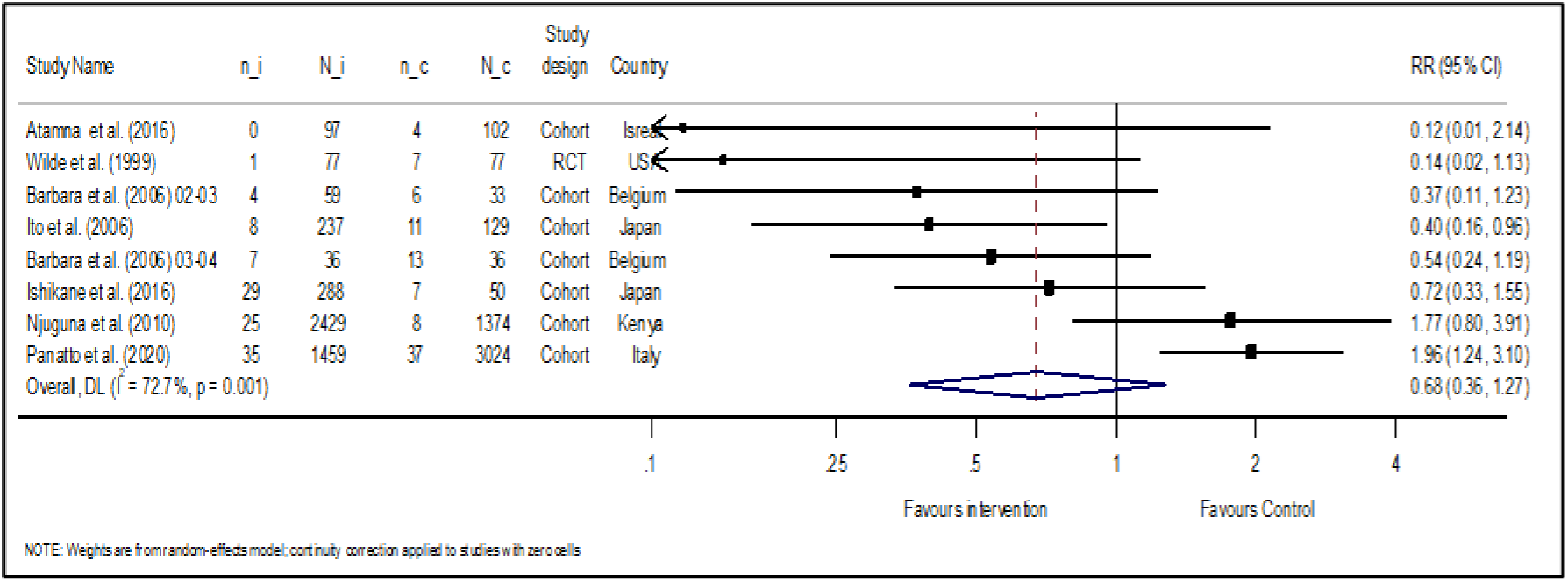
Forest plot of incidence of laboratory-confirmed influenza (LCI) in vaccinated vs. non-vaccinated healthcare workers, n_i: number of HCWs with LCI in the intervention group, N_i: total number of HCWs in the intervention arm, n_c: number of HCWs with LCI in the comparator group, N_c: total number of HCWs in the comparator arm, RR: risk ratio.

**Figure 4.**
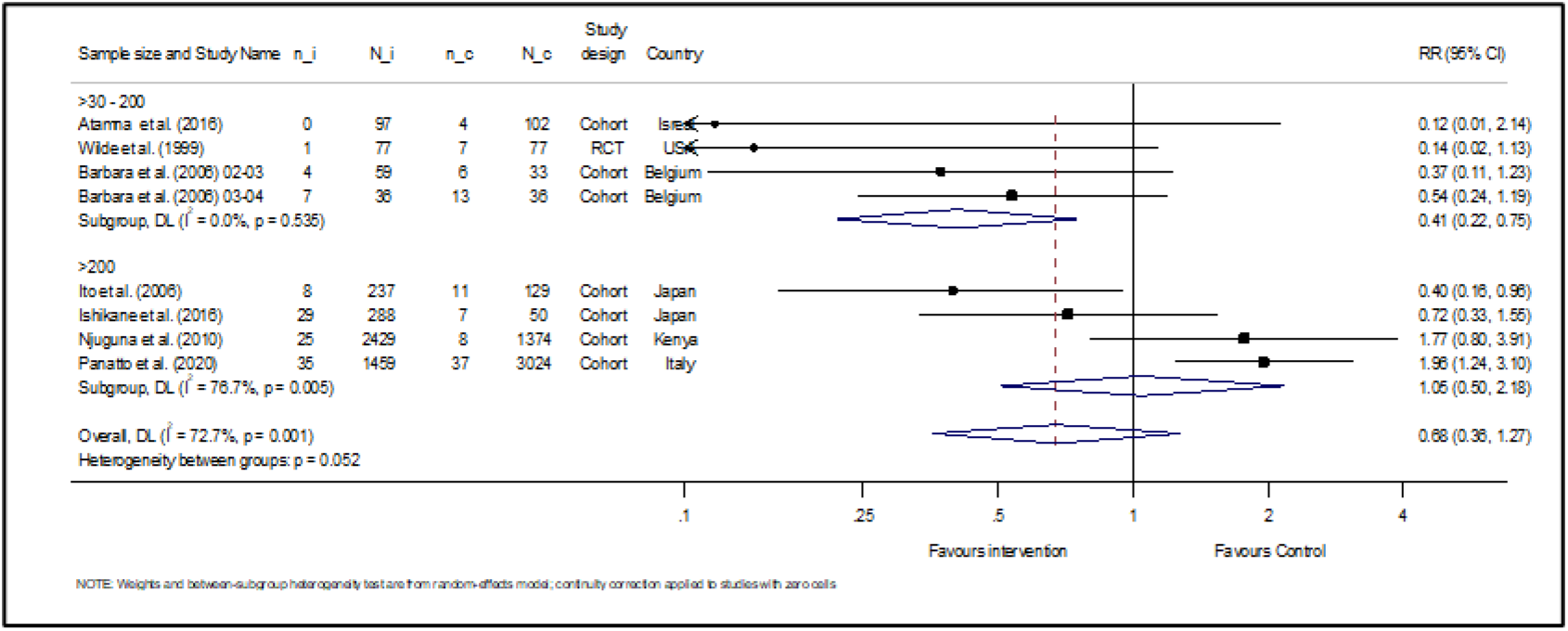
Forest plot of incidence of laboratory-confirmed influenza (LCI) in vaccinated vs. non-vaccinated healthcare workers sub-grouped by study size, n_i: number of HCWs with LCI in the intervention group, N_i: total number of HCWs in the intervention arm, n_c: number of HCWs with LCI in the comparator group, N_c: total number of HCWs in the comparator arm, RR: risk ratio.

In the sensitivity analysis, the observed non-significant reduction in the risk of LCI was retained in the four cohort studies after 2010 (pooled RR: 1.24, 95% CI: 0.62–2.46) and the three studies after 2015 (pooled RR: 0.96, 95% CI: 0.33–2.78). Nevertheless, this demonstrates the robustness and stability of our analysis.

#### Face mask intervention– Laboratory-Confirmed Influenza (LCI)

Three studies reported influenza infections in HCWs, confirmed by laboratory testing for face masks (Table 1), one RCT (MacIntyre et al., 2015), one cohort study (Jaeger, 2011) and one cross-sectional study (Toyokawa, 2011).

The analysis shows a statistically non-significant reduction in the incidence of laboratory-confirmed influenza among HCWs using masks compared to non-users by 77% (pooled RR: 0.23, 95% CI: 0.03–1.68). The included studies show substantial heterogeneity (I^2^: 66.9%, *p*-value = 0.049).

#### Influenza vaccine – Influenza-like illness

Twelve studies compared the differences in the incidence of ILI between the vaccinated and control groups of HCWs (Table 1); two RCTs: (Weingarten, 1988) and (Saxen, 1999), eight cohort studies: (Chan A. L., 2008), (Kheok, 2008), (Igari, 2011), (Njuguna, 2013), (Atamna, 2016), (Panatto, 2020), (Barbara, 2006) and (Fujita, 2009) with 2-year data in the latter two studies and two cross-sectional studies: (Lester, 2003) and (Ng, 2009). The studies were conducted in North America, Europe, Africa, the Middle East and Far East Asia. The pooled effects using a DerSimonian and Laird model shows a statistically insignificant effect size between the vaccinated groups and the comparison groups (pooled RR= 1.04, 95% CI: 0.84 to 1.29, *p*-value= 0.734 (Figure 5).

**Figure 5.**
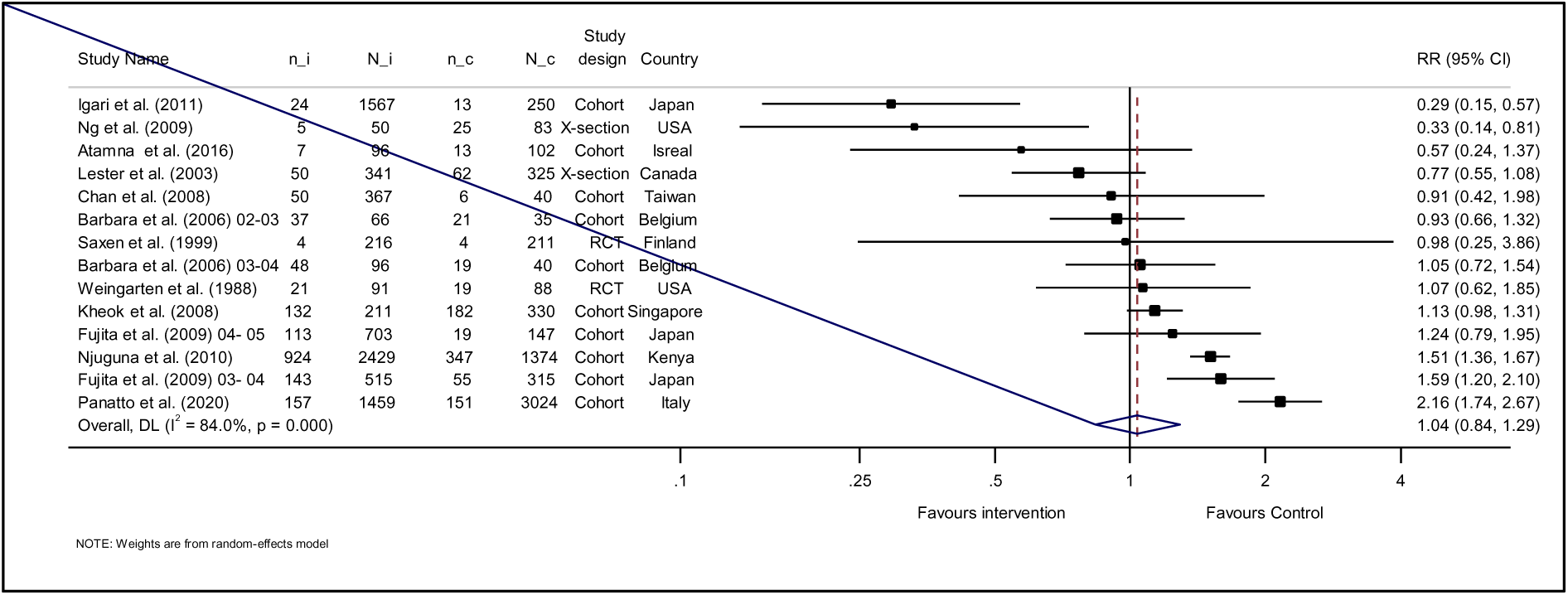
Forest plot of the incidence of Influenza-like illness (ILI) among vaccinated vs. non-vaccinated healthcare workers, n_i: number of HCWs with LCI in the intervention group, N_i: total number of HCWs in the intervention arm, n_c: number of HCWs with LCI in the comparator group, N_c: total number of HCWs in the comparator arm, RR: risk ratio.

The pooled analysis showed considerable heterogeneity between the studies I^2^= 84.0%: *p*-value < 0.001). The subgroups of cohort studies and large studies (i.e. > 200 participants), which contained the most articles, showed statistically non-significant pooling effects (pool effect= 1.15, 95% CI: 0.91-1.44, *p*-value = 0.242) and (pool effect= 1.16, 95% CI: 0.90-1.50, *p*-value = 0.254), respectively. Both subgroups were considerably heterogeneous (I^2^= 85.1%, *p*-value < 0.001 and I^2^= 87.0% and *p*-value < 0.001, respectively).

#### Meta-regression

We conducted a meta-regression on several covariates associated with the incidence of influenza-like illness. Although large and cohort studies had relatively higher effects in predicting ILI, no statistically significant effect was found for the incidence of ILI (Table 3).

**Table 3.** Random-effects model, regression results for influenza-like illness outcome associated with influenza vaccine studies.

| **Variable** | **Coefficient** | **95% conf. interval** |  | **p-value** |
| --- | --- | --- | --- | --- |
| Study design | -0.294 | -0.798 | 0.210 | 0.220 |
| Sample size | -0.401 | -1.234 | 0.432 | 0.305 |
| Country | -0.019 | -0.138 | 0.100 | 0.721 |
| Year | -0.014 | -0.073 | 0.044 | 0.592 |

#### Influenza vaccine – Days lost of work

A total of seven (7) studies reported on absenteeism in healthcare workers in both intervention arms (Table 1); two RCTs (Weingarten, 1988) and (Wilde JA, 1999), four cohort studies: (Chan et al., 2007), (Chan et al., 2008), (Kheok, 2008) and (Zaffina, 2019), with 2-year data available in the latter study, and one cross-sectional study (Amodio et al. 2010). The studies were conducted in North America, Europe and Far East Asia.

The synthesis of the results showed that the average number of days lost from work was in favour of HCWs who were exposed to influenza vaccination (RR= 0.87, 95% CI: 0.80 to 0.93, *p-*value < 0.001) (Figure 6).

**Figure 6.**
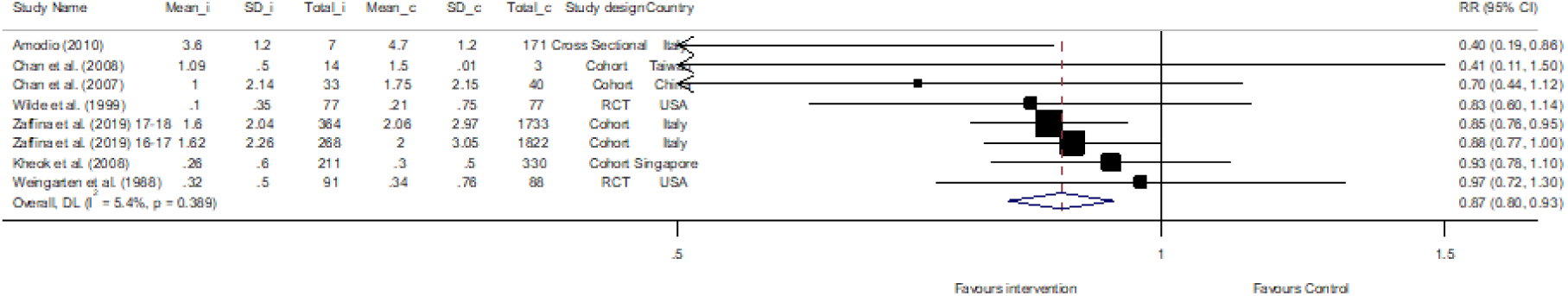
Forest plot of the incidence of absenteeism from work among vaccinated vs. non-vaccianted healthcare workers, Mean_i: events mean in the intervention arm, SD_i: standard deviation of the events in intervention arm, Total_i: total number of participants in the intervention arm, Mean_c: events mean in the comparator arm, SD_c: standard deviation of the events the comparator arm, Total_c: total number of participants the comparator arm, RR: risk ratio.

### Publication Bias

The funnel plots for the laboratory-confirmed influenza and ILI results show asymmetry. The Egger test confirms some bias in the studies (t= −3.30, p= 0.016 and t= −2.45, *p*-value = 0.031, respectively). However, the publication bias was less pronounced in the ILI, as the test power was correspondingly low due to the lower number of participating LCI studies (Figures 7).

**Figure 7.**
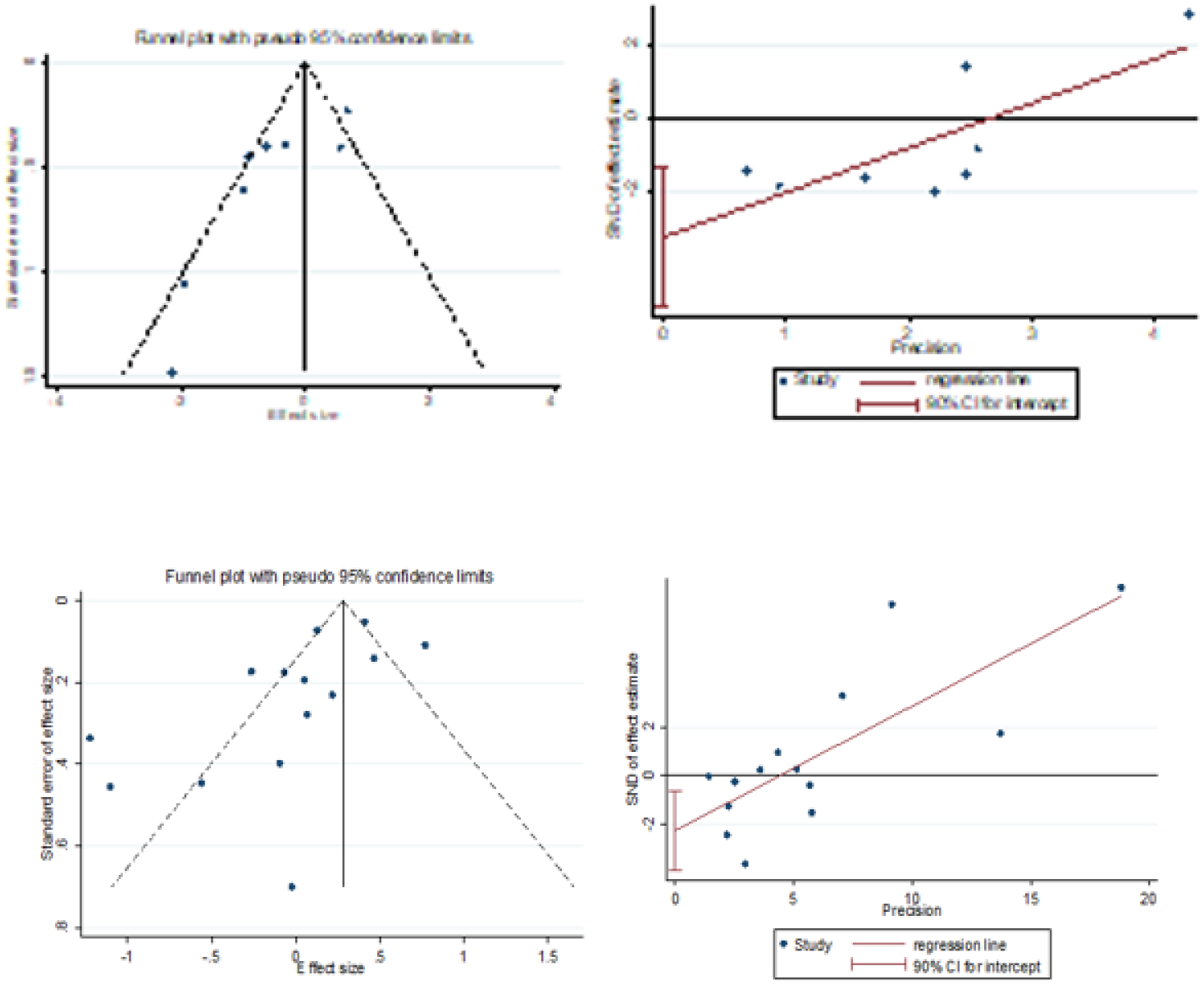
Top row: Funnel plot and Egger test of influenza vaccine and risk of laboratory-confirmed influenza. Bottom row: Funnel plot and Egger test of influenza vaccine and risk of influenza-like illness.

## Discussion

To the best of the authors’ knowledge, this is the most comprehensive review that answers the question of whether the use of different influenza prevention measures (e.g., influenza vaccination or face masks) by healthcare workers have a protective effect against laboratory-confirmed influenza, the incidence of influenza-like illness (ILI), or their work productivity. Our findings do not provide conclusive evidence that vaccination of healthcare workers significantly affects the incidence of laboratory-confirmed influenza (pooled RR: 0.68, 95% CI: 0.36 to 1.27). This result contrasts with what Li et al. (2021) found in their meta-analysis using a similar approach to our study. Li’s smaller sample size compared to our analysis (i.e. 1426 vs. 9507 participants) and the fact that they compared influenza vaccination with meningococcal or pneumococcal vaccination, as in the study by Wilde et al., are the main reasons for this contradiction. On the other hand, our results support the findings of Thomas RE et al. who found no evidence of the benefit of vaccinating healthcare workers for laboratory-confirmed influenza or its complications in people over 60 living in care facilities (14). Although this is a different population group, they represent a high-risk group for infection in a similar setting. The lack of significant findings between vaccinated and unvaccinated healthcare workers may be due to modest vaccine efficacy, variable exposure risks, high use of overlapping protective measures or methodological limitations, such as underpowered studies.

It is interesting to note the different estimates of the effect of influenza vaccination in different subgroups for laboratory-confirmed influenza. For the medium-sized studies, a statistically significant reduction in laboratory-confirmed influenza was demonstrated (pooled RR: 0.41, 95% CI: 0.22 to 0.75; p-value =0.004). The subgroup of large studies clearly shows the influence of the two outliers (i.e. Panatto et al. and Njuguna et al.) on the pooled data. This also reflects the considerable heterogeneity of the studies (I^2^=72.7%, p-value= 0.001), which appears to be due to antigenic mismatch between the circulating strains and the vaccine strains (42) or to, different seasonality in countries with temperate climates (43).

Similar to Li et al. (3754 participants), this review (14571) does not provide sufficient evidence that vaccination of healthcare workers significantly reduces the incidence of influenza-like illness (ILI). The effects were also insignificant in the subgroup and sensitivity analyses compared to Li et al. This was expected as ILI symptoms have a low predictive value (0.30) and low sensitivity (0.27) for influenza infection (44). Pathogens other than the influenza virus can also cause ILI symptoms, which gives reason to consider their role in determining the effect of influenza vaccination in a given year. These reasons make ILI symptoms an unreliable indicator for evaluating the effectiveness of influenza vaccination.

However, our meta-analysis (5329 participants) shows that influenza vaccination reduces the average number of days of absence per vaccinated healthcare worker by 17% compared to non-vaccinated healthcare workers. These findings are consistent with the data from the review by Li et al. (1500 participants), which found a significant reduction in days of absence among vaccinated healthcare workers (summarized SMD= −0.18, 95% CI: −0.28 to −0.07, I^2^ = 28.0%; p = 0.001). These findings indicate that hospitals can save costs overall due to the reduced absenteeism of vaccinated healthcare staff. Nevertheless, these results may not correlate directly with days missed due to influenza-related infection, as they may reflect the complexity of the relationship between vaccination and absenteeism. Absenteeism could be caused by other pathogens that cause ILI symptoms in addition to influenza viruses (45). In addition, other individual factors such as the severity of symptoms, health awareness and the work and living environment could explain this phenomenon (46).

The review also found that in the three studies (1188 participants) involving healthcare workers, there was no statistically significant evidence that face masks reduced the incidence of influenza infection. This is in contrast to the findings of Liang et al. (4751 participants, mostly case-control studies), who found in their Meta-analysis that wearing masks by HCWs significantly reduced the risk of infection of all respiratory viruses between the intervention and control groups (OR: 0.20, 95% CI: 0.11 to 0.37) (47).

Compared to vaccines, the use of face masks by healthcare workers has been much less researched. This is due to ethical limitations in study design, the challenges of standardising and monitoring behavioural intervention, the difficulties in ensuring, measuring and reporting compliance, and the need to control environmental and behavioural confounders such as ventilation, crowding, the use of other personal protective equipment or proximity to patients. These factors make designing and conducting such studies more difficult than vaccine studies. Therefore, the influenza vaccine is the primary means of preventing influenza.

It is important to recognise the limitations and challenges of our work, particularly the restriction to English-language publications. Other shortcomings that we should acknowledge include that most studies did not stratify their analyses according to the different subgroups of healthcare workers (i.e. physicians, nurses, laboratory technicians, etc.) who may have different levels of risk for influenza infection. Second, as there are few placebo-controlled trials for licenced vaccines, we considered including cohort and cross-sectional studies in our search strategy to increase the power of our analysis. We also considered all the studies that met our eligibility criteria and were from different geographical areas, regardless of their quality rating. Furthermore, a non-uniform methodological design increases heterogeneity due to different control of confounding factors. These settings would affect validity and contextual relevance regarding staff, patient populations, infection control norms and exposure risk. Pooled estimates can mask the facility-specific effectiveness of interventions. Third, although most healthcare workers wear at least a face mask in practise to prevent virus transmission [48], it would be difficult to separate the effects of influenza vaccination from the confounding effects of face masks in the studies that examined the effects of influenza vaccination on healthcare workers. Despite these challenges, we conducted a thorough literature search to adequately cover the available literature for this study question, which gives us confidence in the thoroughness of our analyses.

Nevertheless, it is essential to emphasise that influenza vaccination of healthcare workers has been shown to be a valuable strategy to reduce influenza transmission (49). Healthcare workers play a central role in patient care, and their vaccination could contribute to a culture of safety and patient protection. As the evidence for a positive effect of influenza vaccination, specifically in healthcare workers, is limited, other protective measures such as hand hygiene, face masks, and the use of other personal protective equipment (PPE) should be taken in parallel as practical approaches. Further studies are needed to evaluate the impact of other measures to protect against influenza in this study group, which is crucial for understanding the pure effect of the influenza vaccine, with the legitimate purpose of assessing the clinical impact of vaccination in its broader contribution to reducing influenza-related morbidity and mortality in vulnerable groups.

### Practical recommendation

While our meta-analysis found no statistically significant protective effect of influenza vaccination or mask-wearing on laboratory-confirmed influenza or ILI, the 20% reduction in lost working days underlines the practical value for workforce resilience and suggests a real benefit for occupational health. Vaccination helps to maintain staffing levels and reduce the operational burden during the flu season. However, the fact that vaccinated healthcare workers lose fewer working days suggests a link between vaccination and reduced severity of illness. This argues in favour of a robust sickness absence policy to prevent presenteeism. Coupled with other evidence-based infection control strategies that focus on high-risk units and high-exposure roles to provide additional physical barriers, such as other personal protective equipment and infection control measures to reduce the risk of droplet infection transmission, particularly during flu season or respiratory disease outbreaks, this protects staff health, ensures continuity of services and improves wellbeing in the workplace.

## Conclusion

In conclusion, these meta-analyses do not provide conclusive evidence of the effectiveness of influenza vaccination in reducing the incidence of laboratory-confirmed influenza or influenza-like illness in healthcare workers. Medium-sized studies show statistical significance in reducing laboratory-confirmed influenza compared to larger studies, likely because of small effect, methodological differences or heterogeneity. On the other hand, influenza vaccination of healthcare workers significantly reduces the number of lost working days by 17%. Due to the few eligible studies, we could not draw definitive conclusions about the effects of face mask use on specific outcomes. Despite the modest results, vaccination and the use of face masks continue to be recommended by major health organisations (CDC, WHO) for healthcare workers. However, the ineffectiveness may not be due to the tools themselves but to poor implementation (e.g. inconsistent use of masks, delayed vaccination). Vaccination and mask use alone do not significantly reduce confirmed infections or ILI incidence, so hospitals should integrate vaccination and mask use into a broader infection control package (e.g. hand hygiene, ventilation, patient cohorts) rather than treating them as stand-alone measures. Infection control policy should fund high-quality RCTs, pilot programmes and continuous improvement programmes to test combinations of these measures and evaluate the ultimate impact of these protective measures on healthcare workers in different clinical settings to optimise interventions based on local data, with the aim of prioritising multimodal infection prevention strategies that integrate environmental, behavioural and procedural controls.

## Data Availability

All data generated or analysed during this study are included in this published article, and its supplementary information files.

## List of abbreviations

CDC: Centers for Disease Control and Prevention.
GISRS: The WHO’s Global Influenza Surveillance and Response System.
HCWs: Healthcare Workers.
IIV: Inactivated Influenza Vaccine.
ILI: Influenza-Like Illness.
LAIV: Live Attenuated Influenza Vaccine.
LCI: Laboratory-Confirmed Influenza.
MD: Mean Difference.
NOS: Newcastle-Ottawa Scale.
PI/ECO: Population, Intervention/Exposure, Comparison, and Outcome.
PRISMA: Preferred Reporting Items for Systematic Reviews and Meta-Analysis.
RCTs: Randomised Controlled Studies.
RIDT: Rapid Influenza Test.
RR: Risk Ratio.
WHO: World Health Organisation.

## Declarations

### Ethics approval and consent to participate

Not applicable

### Clinical trial number

Not applicable.

### Consent for publication

Not applicable

### Authors’ contributions

ST and KS: contributed to the literature search, study selection, data extraction, and quality assessment.

RHA: contributed to the manuscript writing and provided thoughtful reviews for the manuscript revision.

SF: supervised the research and provided thoughtful reviews for the manuscript revision.

AT: conceptualised and designed the study, contributed to the literature search, study selection, data extraction, and quality assessment, performed the synthesis and interpretation of the results and drafted the manuscript.

All authors read and approved the results and final manuscript.

## Acknowledgements

Not applicable

## Authors’ information

AT: Post-graduate, Kellogg College, University of Oxford, UK.

ST and KS: College of Medicine and Health Sciences, United Arab Emirates University, United Arab Emirates.

RHA: Infectious Diseases Epidemiology Research Advancement (IDERA) Unit, Institute of Public Health, College of Medicine and Health Sciences, United Arab Emirates University, United Arab Emirates.

SF: Nuffield Department of Primary Care Health Sciences, University of Oxford, UK.

AT originally conducted a meta-analysis on the title question as part of a dissertation project, which was part of the requirements for a master’s degree in Evidence-Based Healthcare at the University of Oxford, under the supervision of SF.

