## Supplementary appendix for "Effectiveness of Influenza-Prevention Interventions among Healthcare Workers: A Systematic Review and Meta-Analysis of Health Outcomes": Supplementary appendix final....docx

This appendix formed part of the original submission.

A. **Methods Appendix**
   1. A1. PI/ECO (population, intervention/exposure, comparison, and outcome)

**Study Population**

### We included studies if the population was healthcare workers (HCWs) working in hospitals or medical clinics. These include physicians, nurses, emergency medical personnel, dentists and students, medical and nursing students, laboratory technicians, pharmacists, hospital volunteers and administrative staff.

**Intervention/Exposure**

### The studies' inclusion criteria focused on two distinct types of interventions or exposures (pharmaceutical and non-pharmaceutical). The pharmaceutical interventions focused on influenza vaccination, which included both killed and live attenuated vaccines. For the non-pharmaceutical interventions, the wearing of face masks (medical, surgical, or N95 respirators) was the primary intervention studied.

**Comparison**

### The comparison group consisted of studies that included the population that did not receive any pharmaceutical measures or were not exposed to the non-pharmaceutical measures against influenza.

**Outcomes of interest**

### Studies were considered eligible if they addressed the effectiveness of any of the following: (1) Incidence of laboratory-confirmed influenza infections, (2) Incidence of influenza-like illness, and (3) Absenteeism from work.

2. Search strategy

**PUBMED**

*Search date: 30/09/2024*

Results: 801

"Influenza vaccine*" OR "Influenza vaccination*" OR "Flu vaccine*" OR "Flu vaccination*" OR "Flu shot*" OR "Influenza shot*", ("Influenza Vaccines"[Mesh]), "facial mask*" OR "protective mask*" OR "mask protection" OR "facial protection" OR "face protection", "Respiratory Protective Devices"[Mesh], "Health care worker*" OR "Healthcare worker*" OR "Healthcare professional*" OR "Health care professional*", "Health Personnel"[Mesh:NoExp], Effective* OR efficacy OR impact* OR benefit* OR assessment OR evaluation, "Comparative Effectiveness Research"[Mesh], and outcome* OR absenteeism OR infection* OR episode* OR "laboratory-confirmed-infection*", "Patient Outcome Assessment"[MeAsh].

**Scopus**

*Search date: 30/09/2024*

*Results: 194*

(TITLE-ABS-KEY (effective* OR efficac* OR impact* OR outcome* OR benefit*) AND TITLE-ABS-KEY ("protective measures" OR "protective interventions" OR "preventive measures" OR "safety measures") AND TITLE-ABS-KEY ("influenza vaccine" OR "flu vaccine" OR "influenza immunization" OR "flu shot") AND TITLE-ABS-KEY ("face mask" OR "surgical mask" OR respirator OR mask) AND TITLE-ABS-KEY ("healthcare workers" OR "healthcare professionals" OR "medical staff" OR "hospital staff" OR nurses OR doctors) AND TITLE-ABS-KEY ("laboratory confirmed influenza" OR "confirmed influenza" OR "lab confirmed influenza") AND TITLE-ABS-KEY ("influenza like illness" OR "ILI" OR "flu like illness" OR "influenza symptoms") AND TITLE-ABS-KEY ("days lost at work" OR absenteeism OR "work absence" OR "sick leave" OR "lost work days"))

**Google Scholar**

*Search date: 30/09/2024*

*Results: 107*

"Influenza vaccine*" OR "Influenza vaccination*" OR "Flu vaccine*" OR "Flu vaccination*" OR "Flu shot*" OR "Influenza shot*", OR "facial mask*" OR "protective mask*" OR "mask protection" OR "facial protection" OR "face protection", AND "Health care worker*" OR "Healthcare worker*" OR "Healthcare professional*" OR "Health care professional*", AND Effective* OR efficacy OR impact* OR benefit* OR assessment OR evaluation, AND (outcome* OR absenteeism OR infection* OR "laboratory-confirmed-infection*")

B. **Results Appendix**
   1. **References of included studies**

Amodio E, Anastasi G, Di Pasquale M, Gelsomino V, Morici M, Romano N, Torregrossa MV, Cannova L, Calamusa G, Firenze A. Influenza vaccination among healthcare workers and absenteeism from work due to influenza-like illness in a teaching hospital in Palermo. Italian Journal of Public Health. 2010 Sep 30;7(3).

Atamna Z, Chazan B, Nitzan O, Colodner R, Kfir H, Strauss M, Schwartz N, Markel A. Seasonal Influenza Vaccination Effectiveness and Compliance among Hospital Health Care Workers. The Israel Medical Association journal: IMAJ. 2016 Jan 1;18(1):5-9.

| Barbara M, Hilde P, Samuel C, Fernande Y, Toon S, Sofie S, Joke D, Paul VR. The effect of giving influenza vaccination to general practitioners: a controlled trial [NCT00221676]. BMC medicine. 2006 Dec;4:1-0. |
| --- |
| Chan AL, Shie HJ, Lee YJ, Lin SJ. The evaluation of free influenza vaccination in health care workers in a medical center in Taiwan. Pharmacy World & Science. 2008 Jan;30:39-43. |

Chan SS. Does vaccinating ED health care workers against influenza reduce sickness absenteeism?. The American journal of emergency medicine. 2007 Sep 1;25(7):808-11.

Fujita Y, Okada T, Mugishima H, Kumasaka K, Sawa M, Tachihara S. Trial of influenza HA vaccination for healthcare workers in consecutive years. Japanese journal of infectious diseases. 2009 Nov 30;62(6):464-6.

Igari H, Watanabe A, Chiba H, Shoji K, Segawa S, Nakamura Y, Watanabe M, Suzuki K, Sato T. Effectiveness and safety of pandemic influenza A (H1N1) 2009 vaccine in healthcare workers at a university hospital in Japan. Japanese Journal of Infectious Diseases. 2011 May 31;64(3):177-82.

Ishikane M, Kamiya H, Kawabata K, Higashihara M, Sugihara M, Tabuchi A, Kuwabara M, Yahata Y, Yamagishi T, Odagiri T, Sugiki Y. Seasonal influenza vaccine (A/New York/39/2012) effectiveness against influenza A virus of health care workers in a long term care facility attached with the hospital, Japan, 2014/15: A cohort study. Journal of Infection and Chemotherapy. 2016 Nov 1;22(11):777-9.

Ito Y, Kato T, Sumi H. Evaluation of influenza vaccination in health-care workers, using rapid antigen detection test. Journal of infection and chemotherapy. 2006 Jan 1;12(2):70-2.

Jaeger JL, Patel M, Dharan N, Hancock K, Meites E, Mattson C, Gladden M, Sugerman D, Doshi S, Blau D, Harriman K. Transmission of 2009 pandemic influenza A (H1N1) virus among healthcare personnel—Southern California, 2009. Infection Control & Hospital Epidemiology. 2011 Dec;32(12):1149-57.

Kheok SW, Chong CY, McCarthy G, Lim WY, Goh KT, Razak L, Tee NS, Tambyah PA. The efficacy of influenza vaccination in healthcare workers in a tropical setting: a prospective investigator blinded observational study. ANNALS-ACADEMY OF MEDICINE SINGAPORE. 2008 Jun 1;37(6):465.

Lester RT, McGeer A, Tomlinson G, Detsky AS. Use of, effectiveness of, and attitudes regarding influenza vaccine among house staff. Infection Control & Hospital Epidemiology. 2003 Nov;24(11):839-44.

MacIntyre CR, Seale H, Dung TC, Hien NT, Nga PT, Chughtai AA, Rahman B, Dwyer DE, Wang Q. A cluster randomised trial of cloth masks compared with medical masks in healthcare workers. BMJ open. 2015 Apr 1;5(4):e006577.

Ng TC, Lee N, Hui SC, Lai R, Ip M. Preventing healthcare workers from acquiring influenza. Infection Control & Hospital Epidemiology. 2009 Mar;30(3):292-5.

Njuguna H, Ahmed J, Oria PA, Arunga G, Williamson J, Kosgey A, Muthoka P, Mott JA, Breiman RF, Katz MA. Uptake and effectiveness of monovalent influenza A (H1N1) pandemic 2009 vaccine among healthcare personnel in Kenya, 2010. Vaccine. 2013 Sep 23;31(41):4662-7.

Panatto D, Lai PL, Mosca S, Lecini E, Orsi A, Signori A, Castaldi S, Pariani E, Pellegrinelli L, Galli C, Anselmi G. Influenza vaccination in italian healthcare workers (2018–2019 season): Strengths and weaknesses. results of a cohort study in two large italian hospitals. Vaccines. 2020 Mar 5;8(1):119.

Saxen H, Virtanen M. Randomized, placebo-controlled double blind study on the efficacy of influenza immunization on absenteeism of health care workers. The Pediatric infectious disease journal. 1999 Sep 1;18(9):779-83.

Toyokawa T, Sunagawa T, Yahata Y, Ohyama T, Kodama T, Satoh H, Ueno-Yamamoto K, Arai S, Araki K, Odaira F, Tsuchihashi Y. Seroprevalence of antibodies to pandemic (H1N1) 2009 influenza virus among health care workers in two general hospitals after first outbreak in Kobe, Japan. Journal of Infection. 2011 Oct 1;63(4):281-7.

Weingarten S, Staniloff H, Ault M, Miles P, Bamberger M, Meyer RD. Do hospital employees benefit from the influenza vaccine? A placebo-controlled clinical trial. Journal of General Internal Medicine. 1988 Jan;3:32-7.

Wilde JA, McMillan JA, Serwint J, Butta J, O'Riordan MA, Steinhoff MC. Effectiveness of influenza vaccine in health care professionals: a randomized trial. Jama. 1999 Mar 10;281(10):908-13.

Zaffina S, Gilardi F, Rizzo C, Sannino S, Brugaletta R, Santoro A, Castelli Gattinara G, Ciofi degli Atti ML, Raponi M, Vinci MR. Seasonal influenza vaccination and absenteeism in health-care workers in two subsequent influenza seasons (2016/17 and 2017/18) in an Italian pediatric hospital. Expert Review of Vaccines. 2019 Apr 3;18(4):411-8.

2. List of excluded studies with reasons

**Table B2.** List of excluded studies with reasons

| **No.** | **Study name** | **Reason for exclusion** | **Citation** |
| --- | --- | --- | --- |
| 1 | Colombo et al. (2006) | Different setting | Not a hospital |
| 2 | Susa et al. (2000) | Non-English study |  |
| 3 | Liu et al (2005) | Non-English study |  |
| 4 | Hui et al (2008) | Different setting | Faculty of dentistry in a university |
| 5 | Nishi et al (2001) | Non-English study |  |
| 6 | Murti et al. (2019) | Inapplicable study design | Case-control study |
| 7 | Rizzuto et al. (2006) | Different setting | Italian MoH |
| 8 | Al Qahtani et al. (2021) | Inapplicable study design | Case-control study |
| 9 | Keitel et al. (1997) | Different population | Not HCWs |
| 10 | Jones (1999) | Incomplete data | No SD |
| 11 | Mitchell et al. (1999) | Incomplete data | Data can’t be extracted |
| 12 | Mostow et al. (1977) | Non-comparable interventions | compared split to whole influenza vaccine |
| 13 | Harada et al. (2003) | Non-English study |  |
| 14 | Loeb et al. (2009) | Non-comparable interventions | compared surgical mask to N95 |
| 15 | Maclntyre et al. (2015) | Non-comparable interventions | compared cloth masks to medical masks |
| 16 | Maclntyre et al. (2014) | Other | Not influenza virus specifically |
| 17 | Ritsuko et al. (2001) | Non-English study |  |
| 18 | Gianino et al. (2017) | Incomplete data | No SD |
| 19 | Costa et al. (2012) | Other | two influenza vaccines in a raw in a pandemic season |
| 20 | Cheng et al. (2010) | Different population | data include merged data for HCWs and patient's |
| 21 | Jacobs et al (2009) | Incomplete data | Incomplete data, definition of ILI is lacking |

4. Meta-analyses forest plots

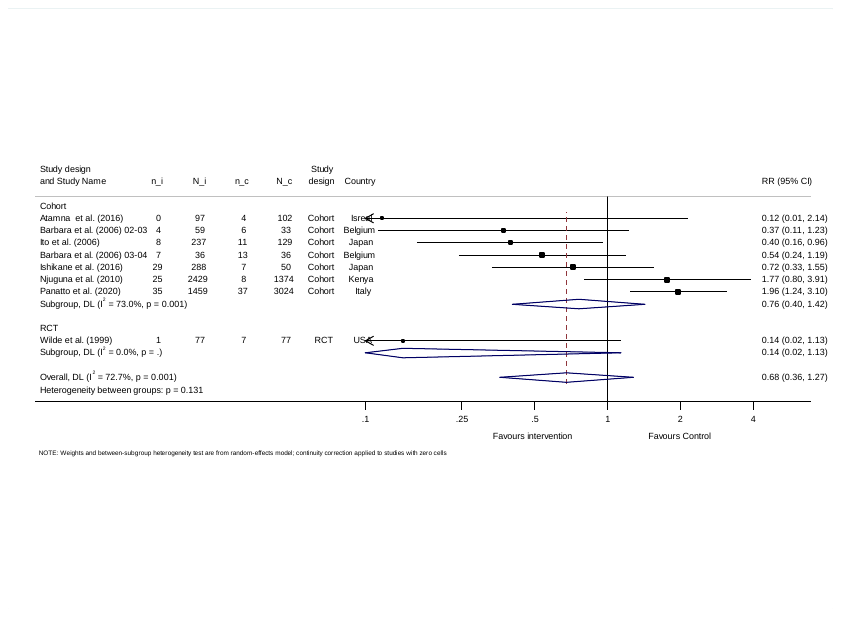

Figure 1. Forest plot of incidence of laboratory-confirmed influenza (LCI) in vaccinated vs. non-vaccinated healthcare workers sub-grouped by study design, n_i: number of HCWs with LCI in the intervention group, N_i: total number of HCWs in the intervention arm, n_c: number of HCWs with LCI in the comparator group, N_c: total number of HCWs in the comparator arm, RR: risk ratio.

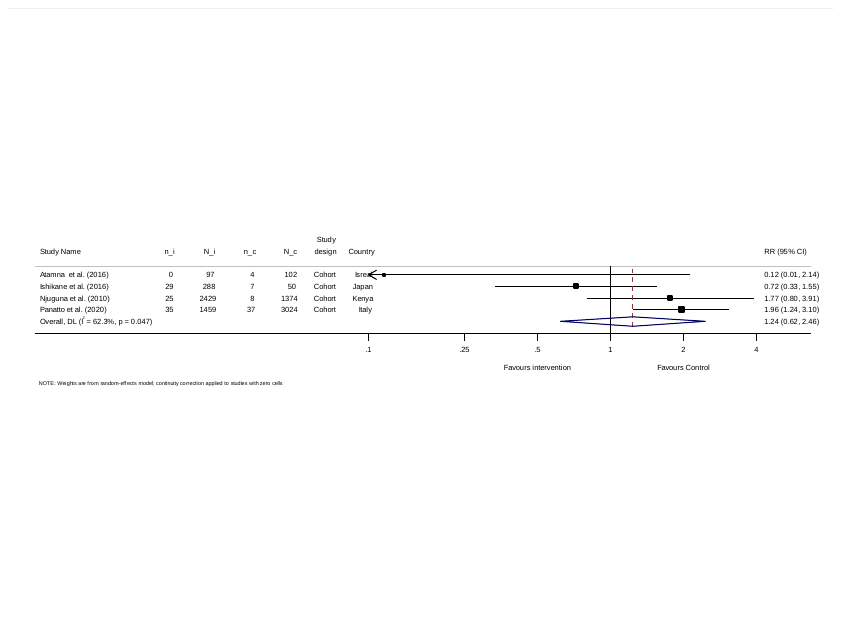

Figure 2. Forest plot of incidence of laboratory-confirmed influenza (LCI) in vaccinated vs. non-vaccinated healthcare workers after removing studies prior to 2010, n_i: number of HCWs with LCI in the intervention group, N_i: total number of HCWs in the intervention arm, n_c: number of HCWs with LCI in the comparator group, N_c: total number of HCWs in the comparator arm, RR: risk ratio.

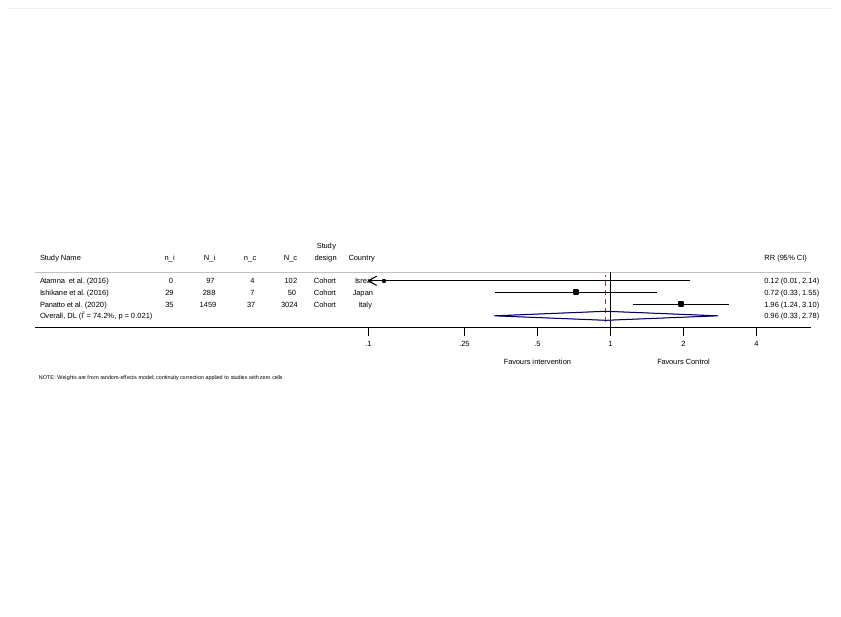

Figure 3. Forest plot of incidence of laboratory-confirmed influenza (LCI) in vaccinated vs. non-vaccinated healthcare workers after removing studies prior to 2015, n_i: number of HCWs with LCI in the intervention group, N_i: total number of HCWs in the intervention arm, n_c: number of HCWs with LCI in the comparator group, N_c: total number of HCWs in the comparator arm, RR: risk ratio.

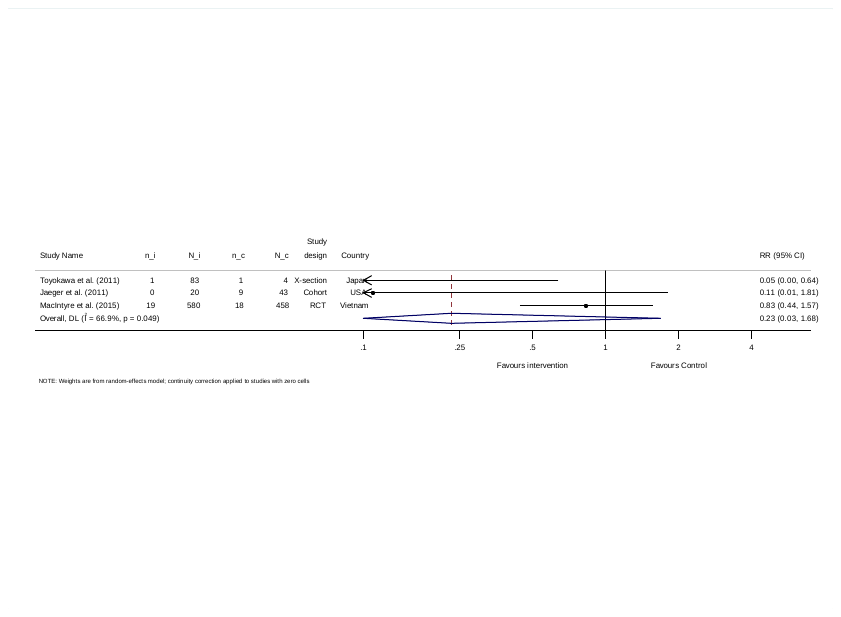

Figure 4. Forest plot of incidence of laboratory-confirmed influenza (LCI) in the mask-users vs. non-mask-users healthcare workers, n_i: number of HCWs with LCI in the intervention group, N_i: total number of HCWs in the intervention arm, n_c: number of HCWs with LCI in the comparator group, N_c: total number of HCWs in the comparator arm, RR: risk ratio.

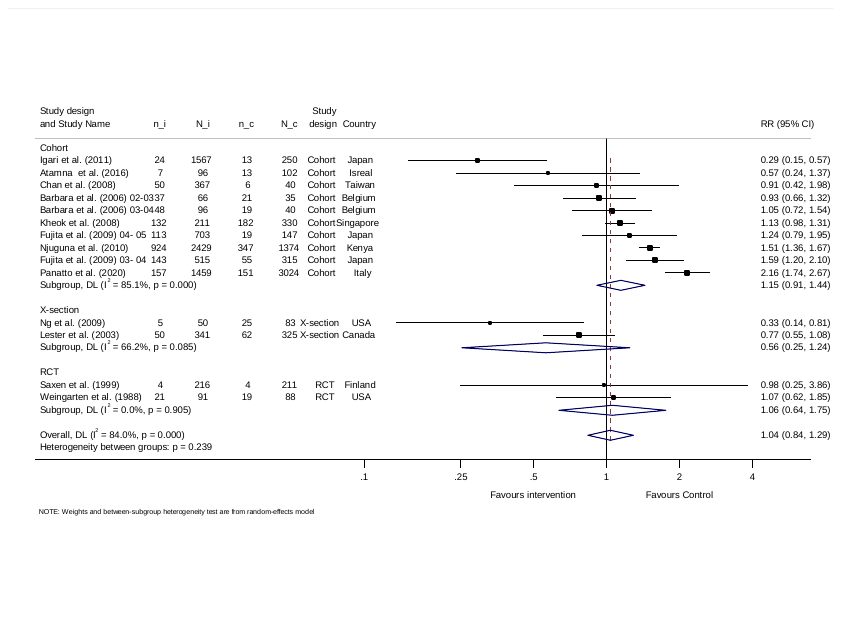

Figure 5. Forest plot of the incidence of influenza-like illness (ILI) among vaccinated vs. non-vaccinated healthcare workers sub-grouped by study type, n_i: number of HCWs with ILI in the intervention group, N_i: total number of HCWs in the intervention arm, n_c: number of HCWs with ILI in the comparator group, N_c: total number of HCWs in the comparator arm, RR: risk ratio.

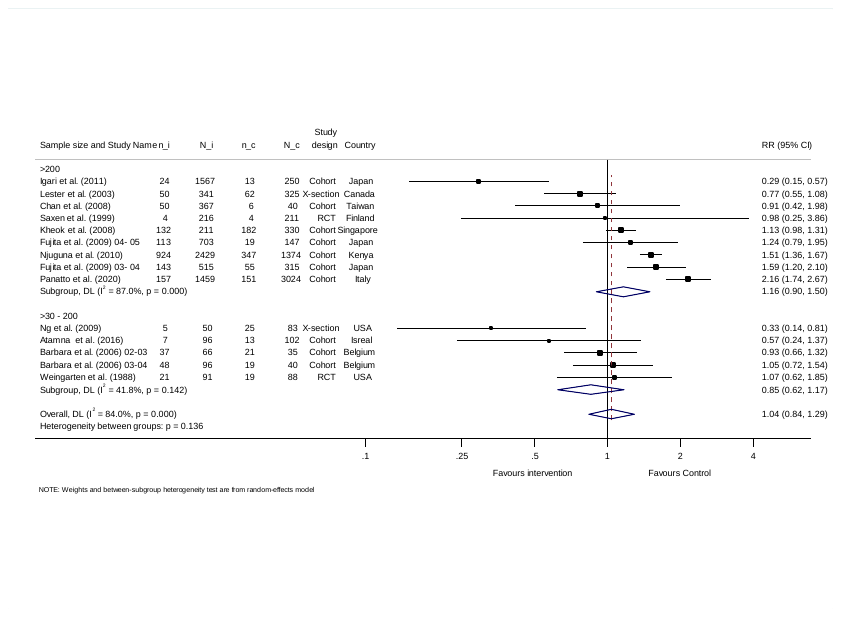

Figure 6. Forest plot of the incidence of incidence influenza-like illness (ILI) among vaccinated vs. non-vaccinated healthcare workers sub-grouped by study sample size, n_i: number of HCWs with ILI in the intervention group, N_i: total number of HCWs in the intervention arm, n_c: number of HCWs with ILI in the comparator group, N_c: total number of HCWs in the comparator arm, RR: risk ratio.

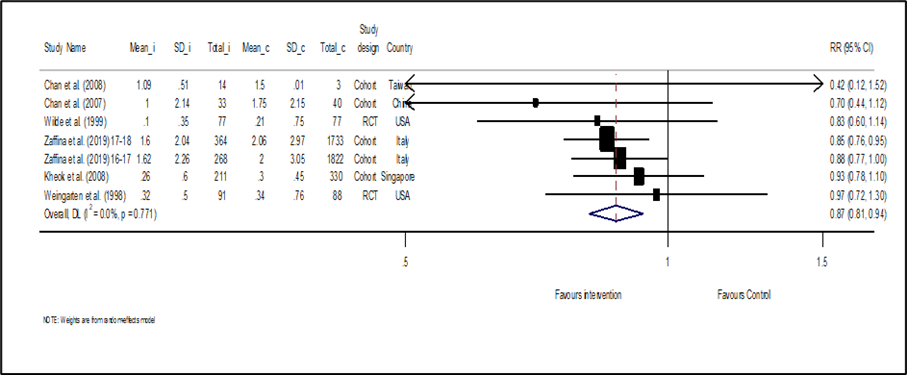

Figure 7. Forest plot of the incidence of absenteeism from work among vaccinated vs. non-vaccianted healthcare workers, Mean_i: events mean in the intervention arm, SD_i: standard deviation of the events in intervention arm, Total_i: total number of participants in the intervention arm, Mean_c: events mean in the comparator arm, SD_c: standard deviation of the events the comparator arm, Total_c: total number of participants the comparator arm, RR: risk ratio.

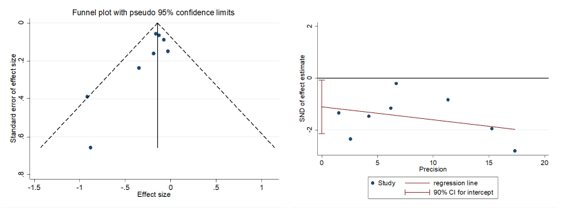

Figure 7. Funnel plot and Egger test of influenza vaccine and days off work.
