## Supplementary appendix for "Effectiveness of Influenza-Prevention Interventions among Healthcare Workers: A Systematic Review and Meta-Analysis of Health Outcomes": Tables final.docx

**List of Tables**

Table 1. Baseline characteristics of the included studies

| **Author Publication year** | **Study design** | **Country** | **Type of intervention** | **Comparison** | **Sample size,**  **(intervention, control)** | **Measured outcome** |
| --- | --- | --- | --- | --- | --- | --- |
| (Wilde JA, 1999) (21) | RCT | USA | Vaccine | Placebo | 154 (77 ,77) | Laboratory-confirmed influenza  Days absent from work |
| (22) (22)  2002-2003  2003-2004 | Cohort | Belgium | Vaccine | Unvaccinated | 92 (59, 33)  72 (36, 36) | Laboratory-confirmed influenza  Influenza-like illness (ILI)  Laboratory-confirmed influenza  Influenza-like illness (ILI) |
| (23) (23) | Cohort | Kenya | Vaccine | Unvaccinated | 3803 (2429, 1374) | Laboratory-confirmed influenza  Influenza-like illness (ILI) |
| (24) (24) | Cohort | Japan | Vaccine | Unvaccinated | 366 (237, 129) | Laboratory-confirmed influenza |
| (25) (25) | Cohort | Israel | Vaccine | Unvaccinated | 199 (97,102) | Laboratory-confirmed influenza  Influenza-like illness (ILI) |
| (26) (26) | Cohort | Japan | Vaccine | Unvaccinated | 338 (288, 50) | Laboratory-confirmed influenza |
| (27) (27) | Cohort | Italy | Vaccine | Unvaccinated | 4483 (1459, 3024) | Laboratory-confirmed influenza  Influenza-like illness (ILI) |
| (28) (28) | RCT | USA | Vaccine | Unvaccinated | 179 (91, 88) | Influenza-like illness (ILI)  Days absent from work |
| (29) (29) | RCT | Finland | Vaccine | Placebo | 427 (216, 211) | Influenza-like illness (ILI) |
| (30) (30) | Cohort | Taiwan | Vaccine | Unvaccinated | 407 (367, 40) | Influenza-like illness (ILI)  Days absent from work |
| (31) (31) | Cohort | Singapore | Vaccine | Unvaccinated | 541 (211, 330) | Influenza-like illness (ILI)  Days absent from work |
| (32) (32) | Cohort | Japan | Vaccine | Unvaccinated | 1817 (1567, 250) | Influenza-like illness (ILI) |
| (33) (33)  2003- 2004  2004- 2005 | Cohort | Japan | Vaccine | Unvaccinated | 830 (515, 315)  850 (703, 147) | Influenza-like illness (ILI)  Influenza-like illness (ILI) |
| (34) (34) | Cross-sectional | Hong Kong | Vaccine | Unvaccinated | 133 (50, 83) | Influenza-like illness (ILI) |
| (35) (35) | Cross-sectional | Canada | Vaccine | Unvaccinated | 666 (341, 325) | Influenza-like illness (ILI) |
| (36) (36)2016-2017  2017-2018 | Cohort | Italy | Vaccine  Vaccine | Unvaccinated  Unvaccinated | 2090 (268, 1822)  2097 (364, 1733) | Days absent from work.  Days absent from work |
| (37) (37) | Cohort | China | Vaccine | Unvaccinated | 73 (33, 40) | Days absent from work |
| (38) (38) | Cross-sectional | Italy | Vaccine | Unvaccinated | 178 (7,171) | Days absent from work |
| (39) (39) | RCT | Vietnam | Mask | No mask | 1,038 (580, 458) | Laboratory-confirmed influenza  Influenza-like illness (ILI) |
| (40) (40) | Cohort | USA | Mask | Glove use | 63 (20, 43) | Laboratory-confirmed influenza |
| (41) (41) | Cross-sectional | Japan | Mask | No mask | 87 (83,4) | Laboratory-confirmed influenza |

Table 2. Risk of bias summary: review authors' judgments for risk of bias items for each included cohort and cross-sectional study. *, the revised study met the criteria for this item, Vxn: vaccine.

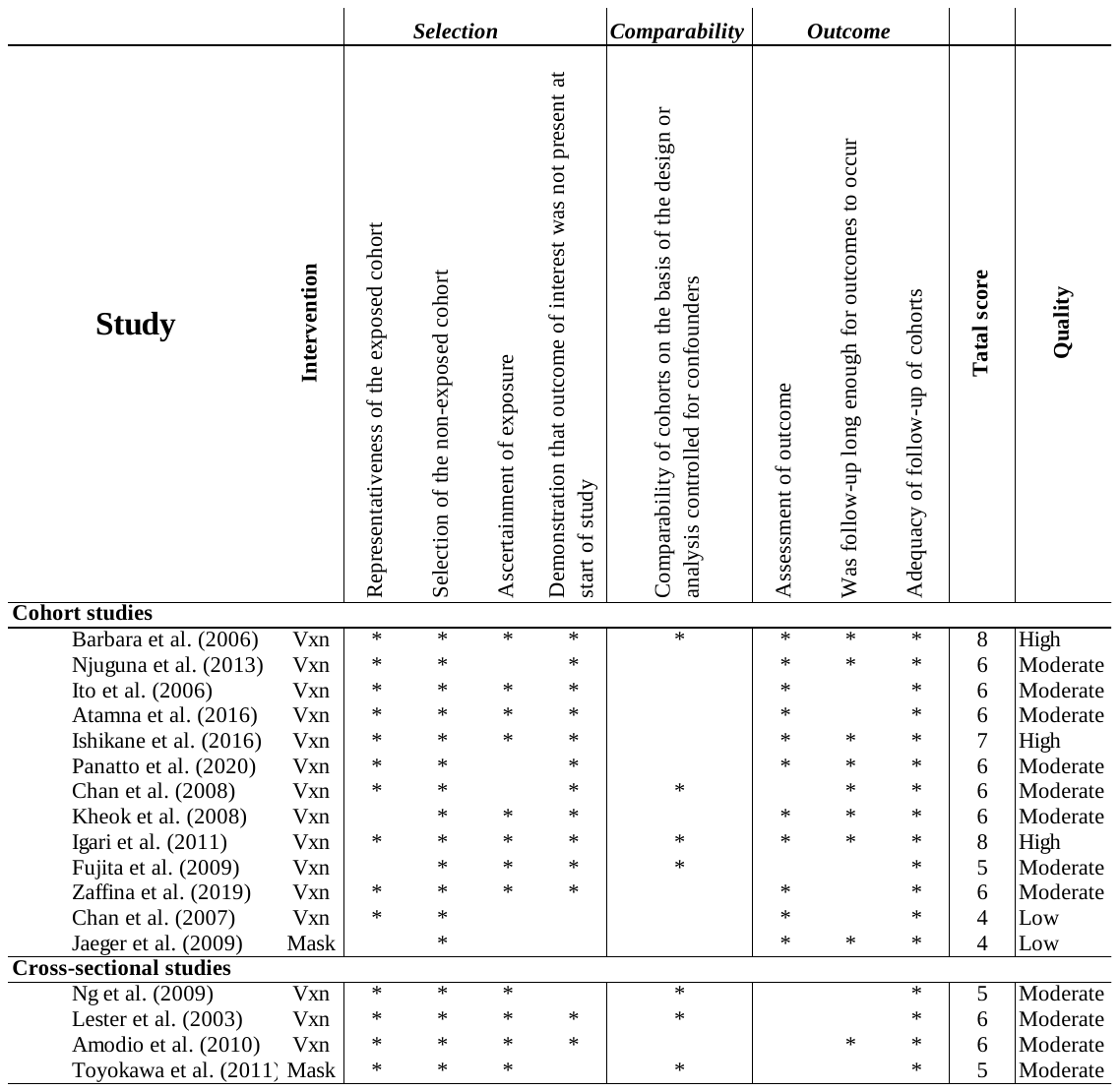

Table 3. Random-effects model, regression results for Influenz-like illness ( ILI) outcome associated with influenza vaccine studies.

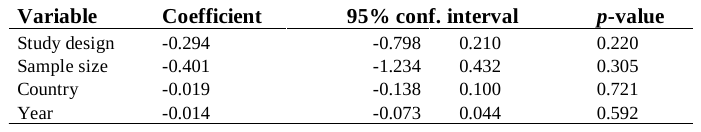
